# The cost-effectiveness of accelerated partner therapy (APT) compared to standard contact tracing for people with chlamydia: an economic evaluation based on the LUSTRUM population-based chlamydia transmission model

**DOI:** 10.1101/2021.07.27.21261128

**Authors:** EV Williams, Ogwulu CB Okeke, CS Estcourt, AR Howarth, A Copas, N Low, C Althaus, F Mapp, Owusu M Woode, M Symonds, TE Roberts

## Abstract

**Objective:** To investigate the cost-effectiveness of accelerated partner therapy (APT) compared with standard contact tracing for people with sexually transmitted chlamydia infection in the United Kingdom

**Design:** Economic evaluation using a model consisting of two components: a population-based chlamydia transmission component, to estimate the impact of APT on chlamydia prevalence, and an economic component, to estimate the impact of APT on healthcare costs and health outcomes.

**Setting:** United Kingdom

**Participants:** Hypothetical heterosexual population of 50,000 men and 50,000 women aged 16-34 years.

**Main Outcome Measures:** Cost-effectiveness based on quality-adjusted life years (QALYs) gained and major outcomes averted (MOA), defined as mild pelvic inflammatory disease (PID), severe PID, chronic pelvic pain, ectopic pregnancy, tubal factor infertility and epididymitis.

**Results:** For a model population of 50,000 men and 50,000 women and an APT intervention lasting 5 years, the intervention cost of APT (£135,201) is greater than the intervention cost of standard contact tracing (£116,334). When the costs of complications arising from chlamydia are considered, the total cost of APT (£370,657) is lower than standard contact tracing (£379,597). Thus, APT yields a total cost saving of approximately £9000 and leads to 73 fewer major outcomes and 21 fewer QALYs lost. Hence, APT is the dominant PN strategy. APT remained cost-effective across the full range of sensitivity analyses.

**Conclusions:** Based on cost-effectiveness grounds APT is likely to be recommended as an alternative to standard contact tracing for chlamydia infection in the United Kingdom

## INTRODUCTION

Sexually transmitted infections (STIs) are a major public health concern and impose a considerable economic burden to healthcare systems such as the UK’s National Health Service (NHS).^1^ Chlamydia infection is the most commonly reported STI in Britain,^2-4^ with 229,411 diagnosed cases in England in 2019.^4^ Treatment and prevention of chlamydia infection is important since untreated infection can progress to long-term complications such as pelvic inflammatory disease (PID), tubal factor infertility (TFI), ectopic pregnancy and chronic pelvic pain in women and epididymitis in men.^5^ However, most chlamydia infections are asymptomatic and remain undiagnosed.^6^ The economic burden of chlamydia infection to the NHS, which is largely attributed to such complications, is estimated at £100 million.^1^

Contact tracing, also referred to as partner notification (PN) is a key element of STI control and involves identifying, testing and treating the sexual partner(s) of individuals with an STI (index patients).^7^ Contact tracing is beneficial to both index patients and their partner(s), and potentially decreases onward transmission within sexual networks and wider populations and reduces reinfection of the index patient from sexual intercourse with an untreated partner.^8^ For chlamydia infection, contact tracing in the UK is typically enhanced patient referral in which a healthcare professional (HCP) advises the index patient to inform their sexual partner(s) of the need for testing and treatment and to refer them to a sexual health service, supplemented with written or website information (hereinafter referred to as standard contact tracing).^7^ Despite substantial resources involved in chlamydia control programmes, these services are performing below expectations.^9,10^ Evidence from the National Chlamydia Screening Programme (NCSP) showed that contact tracing uptake (the number of contacts per index case who were reported as having attended a sexual health service within four working weeks of the date of the contact tracing consultation) decreased from 0.53 in 2016 to 0.42 in 2017 and was below the standard of 0.6.^11^

Studies suggest that expedited partner therapy (EPT), in which the index patient is given antibiotics or a prescription for their sexual partner(s), could be more effective than standard contact tracing and has been widely adopted in the USA.^10 12 13^ However, EPT does not comply with UK prescribing guidance,^14^. We previously developed accelerated partner therapy (APT), a UK-compliant adaption of EPT. In APT a health care professional (HCP) performs a telephone consultation with the sexual partner(s) in private during the index patient’s clinic attendance.^7^ If medically safe, the index patient receives an APT pack, containing antibiotics and self-sampling kits for STIs and HIV to deliver to their sexual partner(s), or the clinic will post the APT pack to the sexual partner(s).^7^

We conducted the *Limiting Undetected Sexually Transmitted infections to RedUce Morbidity* (LUSTRUM) Programme to determine the effectiveness of APT in improving sexual health outcomes in heterosexual men and women. Within the programme, we conducted a cross-over cluster randomised controlled trial (RCT) to compare standard contact tracing for chlamydia infection with APT offered as an additional optional contact tracing method.^7^ The RCT and cost-consequence analysis (CCA) conducted alongside the trial are reported in detail elsewhere.^15^ We conducted the within-trial economic evaluation as a CCA as the trial outcome, cases of chlamydia reinfection avoided, is an intermediate outcome that cannot be used to assess the impact of APT on long term consequences (PID, TFI, ectopic pregnancy, chronic pelvic pain and epididymitis) and their associated costs.^16^ Thus, to better understand the results from the trial and the long-term effects of APT at the population level, we developed a novel deterministic, population-based chlamydia transmission model including the process of contact tracing.^17^ We incorporated the transmission dynamic model output into an economic analysis to estimate the healthcare costs and health outcomes associated with APT and standard contact tracing.

This paper reports the results of the model-informed economic evaluation which compares the cost-effectiveness of APT with standard contact tracing for chlamydia infection. The analysis, which was carried out from an NHS perspective, was based on two outcome measures: cost per major outcomes averted (MOA), which we defined as cases of mild PID, severe PID, ectopic pregnancy, TFI and chronic pelvic pain in women, and epididymitis in men; and cost per quality-adjusted life-year (QALY) gained.

## METHODS

We developed a model to estimate the cost-effectiveness of APT compared with standard contact tracing. Briefly, the model consisted of a transmission dynamic component, required to estimate the impact of APT on chlamydia prevalence, and an economic component, used to estimate the impact of APT on healthcare costs and health outcomes. The analysis was carried out from the perspective of the UK NHS. The outcome measures examined were cost per MOA and cost per QALY gained. We therefore present our results in terms of the incremental cost effectiveness ratio (ICER), namely the additional cost per MOA and additional cost per QALY gained.

### TRANSMISSION DYNAMIC MODEL

We used a deterministic population-based transmission model including a dedicated contact tracing module to estimate the effects of APT on chlamydia prevalence compared with standard contact tracing in Britain. This model has been described in detail elsewhere.^17^ Briefly, the model considered a hypothetical population aged 16-34 and was calibrated to sexual behaviour data between people of the opposite-sex and chlamydia prevalence data reported by 3,671 participants in Britain’s third National Survey of Sexual Attitudes and Lifestyles (Natsal-3, 2010–2012) using Approximate Bayesian Computation (ABC). We simulated the effects of APT on chlamydia transmission by increasing the number of treated partners from current levels by 5%, 10%, 15%, 20%, 25% and 30% respectively.^17^ Based on findings from an exploratory trial of APT, ^18^ our base-case scenario assumed that APT would increase the number of treated partners from current levels by 25%.^18^ The transmission dynamic model predicted that 25% increase in the number of treated partners would reduce chlamydia prevalence by 18% (95% CrI: 5-44%) in both men and women within 5 years.^17^

### ECONOMIC MODEL

We constructed a spreadsheet-based economic model in Microsoft Excel to estimate the number of complications arising from chlamydia infection and their associated healthcare costs and QALYs, and the costs of contact tracing for a hypothetical population of 50,000 men and 50,000 women aged 16-34 after 5 years of APT and standard contact tracing interventions. **Figure 1** below displays a flowchart of the complications we considered in this analysis: mild PID, severe PID, TFI, ectopic pregnancy and chronic pelvic pain in women, and epididymitis in men.

**Fig. 1.**
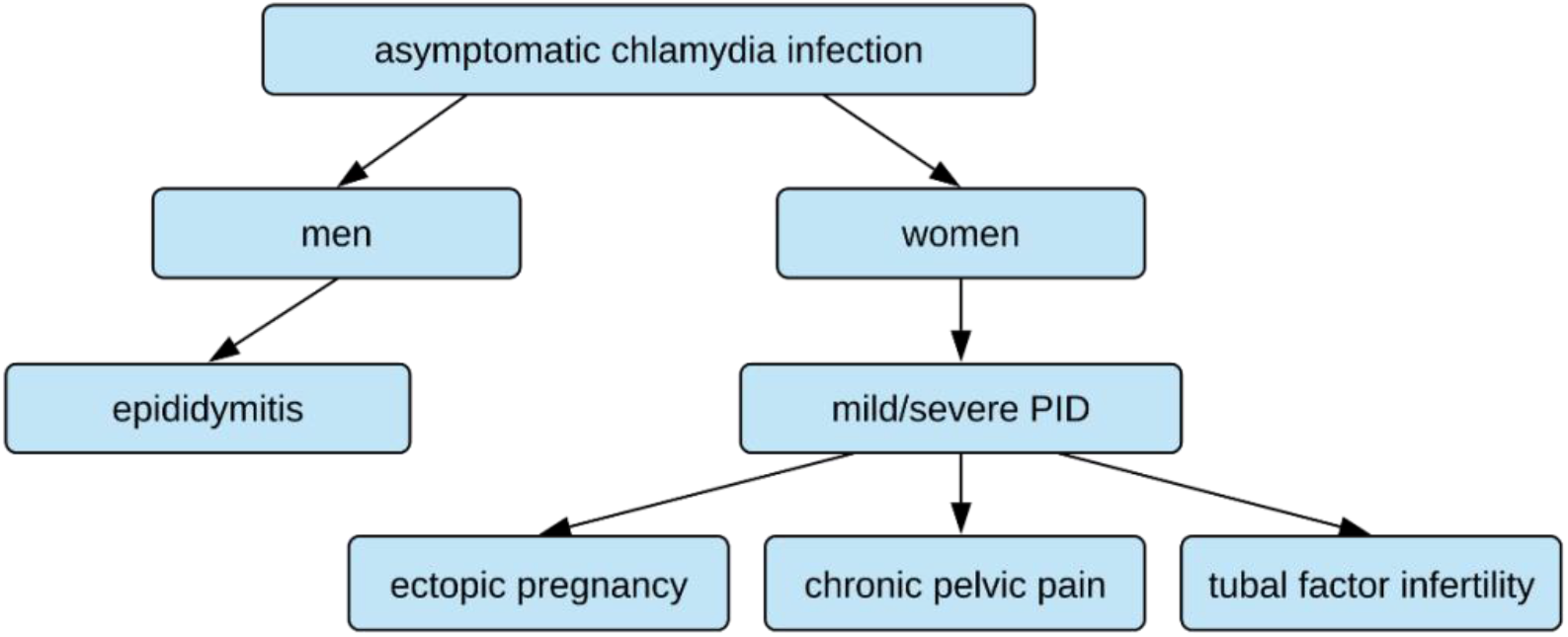
Flowchart of complications arising from chlamydia infection.

For each contact tracing strategy, we used the average output from 1000 simulations of the transmission dynamic model as the base-case to quantify the annual number of index patient (both those notifying and not notifying partners), partners notified (and treated) and incident chlamydia infections within 5 years. We subsequently derived the number of cases of epididymitis in men and PID in women for the stipulated population from the annual number of incident asymptomatic chlamydia infections. We modelled only symptomatic PID as evidence suggests the likelihood of further complications is related to PID symptoms.^19^ We included both mild PID and severe PID in the model: we assumed that 90% of cases were mild, hence were treated in primary care, and 10% of cases were severe and required hospitalisation.^1 20^ We subsequently estimated the number of cases of TFI, ectopic pregnancy and from the modelled cases of PID. We made a simplifying assumption wherein all women considered future pregnancy, thus could experience TFI or ectopic pregnancy. We explored the impact of this assumption in the sensitivity analysis.

Based on these data, we estimated the averted complications (MOAs) from APT compared to standard contact tracing. We additionally incorporated health-state utility values (HSUVs) to estimate QALYs gained. The model analysed the effects of an APT intervention lasting for 5 years compared with standard contact tracing. The time-horizon of the model was 27 years in order to analyse the impact of long-term complications in women (TFI, ectopic pregnancy and chronic pelvic pain).

#### Probabilities

Table 1 shows the probabilities we applied to the economic model to estimate the number of cases of PID and epididymitis progressing from chlamydia infection, and cases of TFI, ectopic pregnancy and chronic pelvic pain arising from PID. We sourced the base-case probabilities from published studies that identified through a review of the literature. We conducted deterministic sensitivity analyses (DSAs) to explore the effect of varying the respective probabilities within an identified range.

**Table 1.** Probabilities applied to the economic model.

| Complication | Probability | Range | Applied to | Source(s) |
| --- | --- | --- | --- | --- |
| PID | 0.100 | 0.02-0.25 | Asymptomatic chlamydia infection (women) | 1,21 22,23,24 |
| → Severe PID | 0.100 | 0.1-0.25 | PID | Assumption based on 1 20 |
| → Mild PID | 0.900 | 0.75-0.9 | PID | Assumption based on 1 20 |
| Ectopic pregnancy | 0.04 | 0.02-0.10 | PID | 19 25-28 |
| TFI | 0.09 | 0.05-0.23 | PID | 25,26-28 |
| Chronic pelvic pain | 0.12 | 0.05 – 0.20 | PID | 19,25,26,27 |
| Epididymitis | 0.02 | 0.01 – 0.04 | Asymptomatic chlamydia infection (men) | 21,29 |

#### Costs and resource use

Table 2 below describes the key costs applied in the economic model. The model included two cost components: the direct costs of the alternative contact tracing strategies and costs of complications arising from chlamydia infection. We derived unit costs from standard data sources, other published sources and the LUSTRUM Programme. All costs sourced are reported in pounds (£) sterling for the 2019/20 price year having been appropriated inflated if necessary. We applied weighted averages where appropriate. We applied the recommended discount rate of 3.5% to future costs and outcomes.^30^ We provide an explanation of how key costs were calculated and assumed resource use for each contact tracing strategy and complication in the Appendix.

**Table 2.** Key costs within the economic model.

| Item | Unit Cost | Source(s) |
| --- | --- | --- |
| *Contract tracing costs* |  |  |
| APT index patient (notifying partner) | £32.09 | 31,32 |
| APT index patient (not notifying partner) | £22.43 | 31,32 |
| APT sex partner | £36.17 | 15 31,32 |
| Standard contact tracing index patient | £22.43 | 31,32 |
| Standard contact tracing sex partner | £38.46 | 15,31,32 |
| *Complications* |  |  |
| Mild PID | £123.15 | 31 32,33,34 |
| Severe PID | £1366.81 | 31 32,33,35 |
| Ectopic pregnancy | £1740.29 | 31 32,33,34,36 |
| TFI | £3639.06 | 31,33,37,38 |
| Chronic pelvic pain | £652.26 | 31,33 |
| Epididymitis | £187.75 | 39 |

#### Outcomes

We examined two outcomes: the number of major outcomes averted (MOA) and quality adjusted life years (QALYs) gained. We defined major outcomes as cases of mild PID, severe PID, TFI, ectopic pregnancy and chronic pelvic pain in women and epididymitis in men. We assessed QALY loss solely for the aforementioned complications, hence, we did not consider the impact of acute chlamydia infection on quality of life.

The HSUVs we used to inform the QALYs are detailed in **Table 3** below.^40^ We previously elicited the HSUVs for mild PID, severe PID, TFI, ectopic pregnancy and chronic pelvic pain from first principles using appropriate conventional and chained time trade-off (TTO) techniques.^40^ They pertain to health states resulting from initial chlamydia infection and are applicable to the UK general population.^40^ No equivalent primary study was conducted for male chlamydia health states, hence we used a previously published HSUV for epididymitis. We additionally utilised health state durations from previously published studies for all complications. We explored the impact of both applying previously published HSUVs for the female complications and varying the health state durations within an identified range using DSAs. We provide further details on the variations applied in the Appendix.

**Table 3.** Health state utility values and durations applied to the economic model.

| Complication | HSUV | Duration (years) | Source(s) |
| --- | --- | --- | --- |
| Mild PID | 0.74 | 0.0274 | <sup>41</sup> |
| Severe PID | 0.76 | 0.0329 | <sup>41</sup> |
| Ectopic pregnancy | 0.46 | 0.0767 | <sup>25,42,39,43,44</sup> |
| TFI | 0.54 | 10 | <sup>21,41,45</sup> |
| Chronic pelvic pain | 0.4 | 5 | <sup>25,41,42,44</sup> |
| Epididymitis | 0.9 | 0.0301 | <sup>39</sup> |

#### Assumptions

We employed several simplifying assumptions to develop a workable model. These are described below:

- In the base-case analysis we assumed that APT increases the number of partners treated from current levels by 25%.
- We assumed that mild PID, severe PID and epididymitis occurred within the year of initial chlamydia infection.^46^
- We assumed that chronic pelvic pain was occurred within 5 years of chlamydia infection.^46^
- We assumed that ectopic pregnancy and the impact of tubal factor infertility occurred 11 and 12 years after initial chlamydia infection respectively. The basis for such assumptions is that the mean age of individuals in the transmission model was 19,^17^ and the mean age of the mother in the UK was reported to be 30 years old.^41^ We assumed that the impact of TFI occurred after 12 months of not becoming pregnant without the use of contraception, thus, TFI modelled to occur one year later than ectopic pregnancy.
- In the base-case analysis we assumed that 10% of PID is mild (treated in primary care) and 90% of PID is severe (required hospitalisation). 1 20
- We assumed that the effects of previous or repeated chlamydia infection and PID do not modify the probability of a woman experiencing further complications.
- We assumed that no complications were incurred during treatment of chlamydia sequalae.
- In the base-case analysis we assumed that that all women considered future pregnancy and that all women experiencing TFI underwent one cycle of in vitro fertilisation (IVF).

#### Analysis

We conducted a cost-effectiveness analysis (CEA) and a cost-utility analysis (CUA) from the perspective of the NHS estimate incremental cost-effectiveness ratios (ICERs). We calculated an ICER as the difference in costs divided by the difference in MOAs/QALYs of the two PN strategies, and present results in terms of the additional cost per MOA and additional cost per QALY gained. For the CUA, we considered the cost-effectiveness of APT in relation to the NICE recommended threshold of £20,000 - £30,000 per QALY gained.

We used sensitivity analyses to explore uncertainty in the model. We undertook a series of one-way/multi-way DSAs to examine the impact of varying key parameters and the assumptions employed by the economic model. These included individually and simultaneously varying all transmission probabilities to upper and lower bounds of the identified range and reducing the APT sex partner consultation time to 6.2 minutes (the mean consultation duration observed in the RCT). The full range of DSAs conducted are reported in the Appendix.

We subsequently conducted scenario analyses where we lowered the increase in the number of partners treated from current levels by APT to 15% and 5% respectively to explore additional scenarios of the impact of APT on contact tracing.

We also conducted probabilistic sensitivity analysis (PSA) to assess parameter uncertainty. Where possible, we assigned distributions to probabilities, HSUVs, and costs within the model. We attached beta distributions were attached to probabilities and HSUVs, and gamma distributions to attached to costs. We used the output from 1000 runs of each year of the transmission dynamic model in the economic model to compute corresponding Monte Carlo simulations, and generated mean cost and effectiveness estimates by simultaneously varying all relevant economic parameters. We used these estimates jointly to form an empirical distribution of the differences in both the cost and effectiveness of interventions. We subsequently generated cost-effectiveness acceptability curves (CEACs) to depict the probabilities that APT is a cost-effective strategy for chlamydia contact tracing compared with standard contact tracing across a range of values representing the decision maker’s willingness to pay (WTP) for an additional benefit.

## Results

### Base-case results

The results of the base-case analysis are shown in **Table 4**. In a model population of 50,000 men and 50,000 women aged between 16-34, the intervention cost of APT was £135,201 whilst the intervention cost of standard contact tracing was £116,334. When the costs of complications arising from chlamydia infection were accounted for the total cost of APT was £370,657 whilst the total cost of standard contact tracing was £379,597. Thus, APT resulted in a total cost saving of £8940. The base-case analysis showed there were 518 major outcomes and 181 QALYs lost from 5 years of APT compared to 591 major outcomes and 202 QALYs lost for standard contact tracing over the same duration. Hence, the results suggest APT is a dominant strategy compared to standard contact tracing (i.e., cost-saving and more effective).

**Table 4.** Base-case results.

| Contact Tracing Approach | Intervention Cost (£) | Total Cost (£) | MOs | QALYs lost | ICER/MOA (£) | ICER/ QALY gained (£) |
| --- | --- | --- | --- | --- | --- | --- |
| APT | 135,201 | 370,657 | 518 | 181 | Dominant | Dominant |
| Standard Contact Tracing | 116,334 | 379,597 | 591 | 202 | - | - |

Note: Costs, MOs and QALYs lost are rounded in the table for presentation. Dominant indicates that strategy is more effective and cost-saving.

### Deterministic sensitivity analysis

We conducted numerous DSAs to examine the impact of varying key parameters and assumptions employed by the model. For the sake of brevity, only the variations that made a substantial difference to the base-case result (progression to PID reduced to 2% and progression to all complications simultaneously reduced to the lowest probabilities in the identified range) are shown in **Table 5**.

**Table 5.** Key deterministic sensitivity analysis results.

| **All complications reduced to lowest probability in range (PID = 2%, TFI = 5%, ectopic pregnancy = 2%, chronic pelvic pain = 5%, epididymitis = 1%)** |  |  |  |  |  |  |
| --- | --- | --- | --- | --- | --- | --- |
| PN Approach | Intervention Cost (£) | Total Cost (£) | MOs | QALYs lost | ICER/MOA (£) | ICER/ QALY gained (£) |
| APT | 135,201 | 173,375 | 113 | 18 | 977 | 6639 |
| Standard Contact Tracing | 116,334 | 159,058 | 128 | 20 | - | - |
| **PID progression reduced to 2%** |  |  |  |  |  |  |
| PN Approach | Intervention Cost (£) | Total Cost (£) | MOs | QALYs lost | ICER/MOA (£) | ICER/ QALY gained (£) |
| APT | 135,201 | 191,040 | 149 | 37 | 593 | 2797 |
| Standard Contact Tracing | 116,334 | 178,862 | 169 | 41 | - | - |
| Note: Costs, MOs, QALYs lost and ICERs are rounded in the table for presentation. |  |  |  |  |  |  |

When we reduced the probability of progression to PID to 2% the total cost of APT decreased to £191,040 whilst the total cost of standard contact tracing decreased to £178,862. Hence, the additional costs of complications arising from chlamydia infection under standard contact tracing no longer exceeded the additional intervention cost for APT. Although APT was no longer cost-saving it still led to fewer major outcomes and QALYs lost. The resultant ICER in terms of QALYs (£2797 per QALY gained) is below the benchmark threshold value of £20,000 per QALY suggesting APT is a cost-effective alternative.

Similarly, when we simultaneously reduced the probabilities of progression to all complications to the lower bounds of the identified range, the total cost of APT (£173,375) exceeded the total cost of standard contact tracing (£159,058). However, APT led to fewer major outcomes and QALYs lost. The resultant ICER (£6639 per QALY gained) remained below the benchmark threshold value, suggesting that APT is cost-effective.

### Scenario Analysis

The results of the scenario analyses in which we lowered the increase in the number of partners treated by APT to 15% and 5% respectively are shown in **Table 6**. Under both scenarios the total cost of APT (£380,157 (*15%*) and £386,426 (*5%*)) was greater than the total cost of standard contact tracing (£379,597) whilst APT led to fewer MOs (542 (*15%*) and 567 (*5%*)) and QALYs lost (189 (15%) and 197 (*5%*)) than standard contact tracing (591 MOs and 202 QALYs). The resultant ICERs in terms of QALYs (£43/QALY gained (*15%*) and £1050/QALY gained (*5%*)) suggest APT is a cost-effective alternative to standard contact tracing.

**Table 6.** Scenario analysis results.

| **APT increases the number of partners treated from current levels by 15%** |  |  |  |  |  |  |
| --- | --- | --- | --- | --- | --- | --- |
| PN Approach | Intervention Cost (£) | Total Cost (£) | MOs | QALYs lost | ICER/MOA (£) | ICER/ QALY gained (£) |
| APT | 132,590 | 380,157 | 542 | 189 | 11 | 43 |
| Standard Contact Tracing | 116,334 | 379,597 | 591 | 202 | - | - |
| **APT increases the number of partners treated from current levels by 5%** |  |  |  |  |  |  |
| PN Approach | Intervention Cost (£) | Total Cost (£) | MOs | QALYs lost | ICER/MOA (£) | ICER/ QALY gained (£) |
| APT | 128,803 | 386,426 | 567 | 197 | 278 | 1050 |
| Standard Contact Tracing | 116,334 | 379,597 | 591 | 202 | - | - |

Note: Costs, MOs, QALYs lost and ICERs are rounded in the table for presentation.

### Probabilistic sensitivity analysis

Figure 2 presents the CEAC between APT and standard contact tracing. The CEAC examines the probability that the PN strategies are cost-effective, compared with the alternative, for a range of values of the maximum acceptable ceiling ratio.^47^ For any possible willingness-to-pay (WTP), the height of each curve indicates the proportion of model replications at which the PN strategy is cost-effective. As the WTP per additional QALY exceeds £250, APT is the preferred PN strategy, with a higher probability of being cost-effective. Given an arbitrary WTP of £2500 per additional QALY the probability that APT is cost-effective compared with standard contact tracing is 95%. The difference in probabilities over the WTP reflects some uncertainty in the model.

## Discussion

### Principal findings

This study used a dynamic transmission model to estimate the cost-effectiveness of APT compared with standard contact tracing. The base-case results suggest that APT is cost-saving and more effective in terms of major outcomes and QALYs, and therefore is a cost-effective alternative to standard contact tracing in the UK context.

**Fig. 2.**
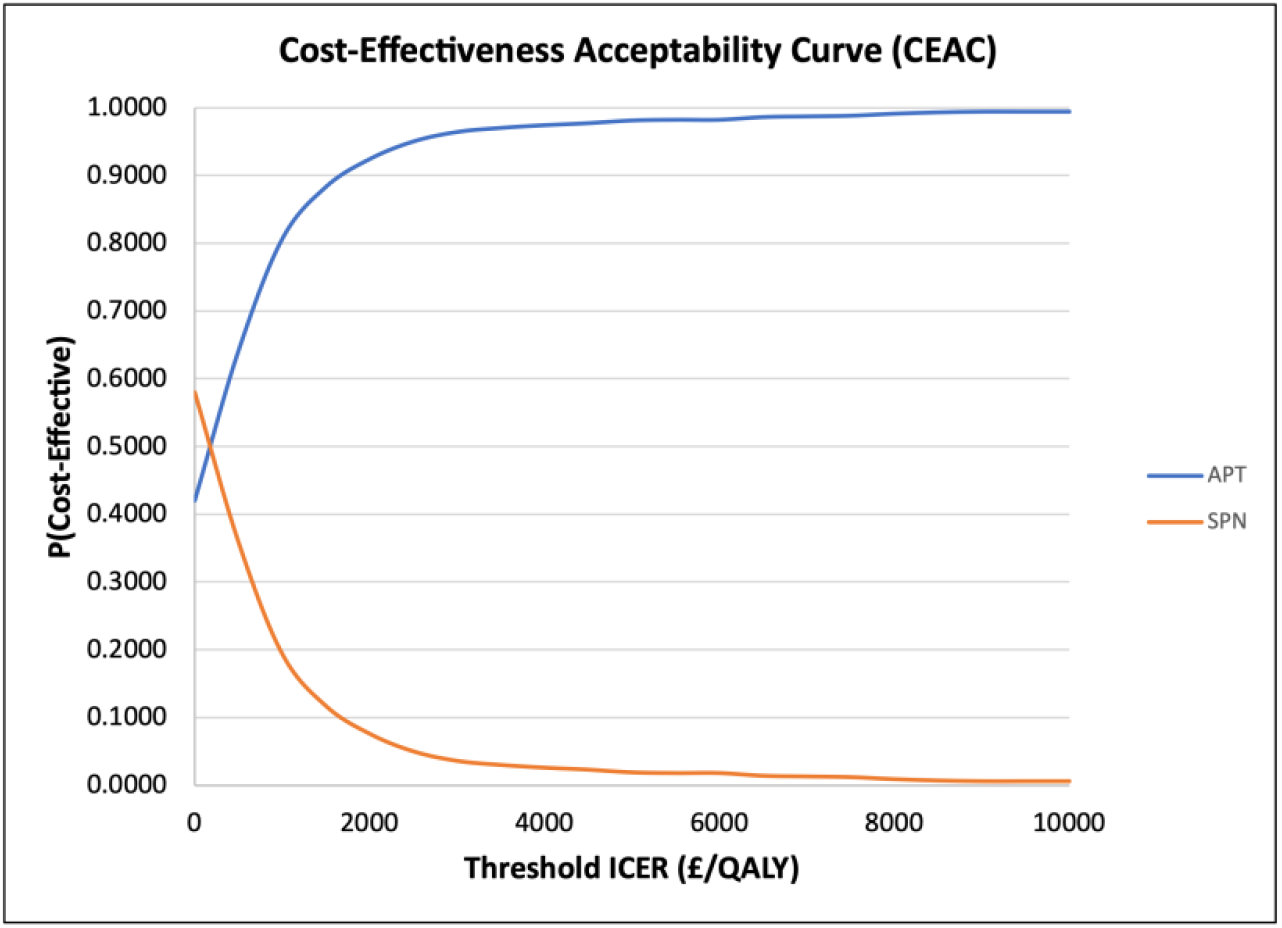
Cost-effectiveness acceptability curve between APT and standard contact tracing, using distributions around the accuracy data.

Previous research has shown that the probability of progression from chlamydia infection to PID is a key parameter when estimating the cost-effectiveness of chlamydia control interventions. Our sensitivity analyses showed that when we varied progression to PID to 2% and 25% respectively APT was a cost-effective, but not cost-saving, alternative. In addition, although the extent to which APT increases the number of treated partners from current levels is uncertain, our scenario analysis suggests that APT is cost-effective for an increase as low as 5%. APT was further shown to remain cost-effective across the full range of DSAs. The PSA reflected these findings and indicated high probability of APT being cost-effective.

### Strengths and limitations

A key strength of our study is that we estimated the cost-effectiveness of APT using a deterministic population-based transmission model, hence, appropriately accounted for dynamic processes such as chlamydial transmission and the effect of APT on chlamydia prevalence.

In addition, this is the first economic evaluation of a chlamydia control intervention, to our knowledge, to use QALYs estimates derived from reliable HSUVs (for the female complications), which were elicited using appropriate methods and relevant to the population under consideration. The model-based results can consequently be used to straightforwardly inform decisions about chlamydia control in the context of limited healthcare resources. Previous economic evaluations of chlamydia control interventions typically relied on HUSVs derived using inadequate methods and for complications not explicitly arising from initial chlamydia infection, ^e.g.22,25,39,42^ thus, substantial uncertainty was associated with their results.

A limitation of our study is that we cautiously used a HSUV estimate from published literature for epididymitis as a robust HSUVs was not available. Our sensitivity analysis showed that varying the epididymitis HSUV had no substantial impact on the results. In addition, there is considerable uncertainty associated with the QALY durations, timings and probabilities of complications, in particular for women, as contraception use can delay their diagnosis for many years and chlamydia is not the only cause. We conducted sensitivity analyses to explore the impact of these uncertain parameters on the cost-effectiveness results and found no substantial impact.

Furthermore, as a necessary simplification of the transmission model, we only considered notification of the index patients’ most recent sex partners.^17^ Given that the average number of partners notified is typically below one, not including these additional partners is unlikely to substantially affect the cost-effectiveness results. Furthermore, we did not consider different age groups, men having sex with men (MSM) and different ethnic groups. As chlamydia infection occurs mostly in the population of young opposite-sex partners, these omissions are unlikely to significantly affect the cost-effectiveness results.

Our study did not consider the effect that repeated or persistent chlamydia or PID would have on tubal damage, thus, the associated results should be interpreted with caution. Finally, we made no comparisons for different forms of APT. A previous cost-consequence analysis considered the costs and effects of APT Hotline, equivalent to the APT intervention modelled by our study and APT Pharmacy, where the sex partner is assessed by a trained community pharmacist.^48^ Future research could assess the cost-effectiveness and patient preferences for different forms of APT.

### Comparison with the literature

To our knowledge, this is the first economic evaluation of APT in terms of cost per MOA and QALY gained. Previous economic evaluations of APT interventions have focused on intermediate outcomes, such as cost per case of reinfection avoided, and are compared to the results of the cost-consequence analysis of the LUSTRUM RCT.^15^

A US-based study compared the cost-effectiveness of EPT,^49^ and standard partner referral (SR) for the treatment of chlamydia or gonorrhoea infections. The study, which was based on two clinical trials, additionally incorporated data on costs of complications (PID, chronic pelvic pain, ectopic pregnancy and TFI) from the literature and previously published HSUVs for cases of PID. The results showed that from a healthcare perspective, EPT was a cost saving alternative to SR ($399.88 compared to $453.17 per male index patient and $168.53 compared to $194.25 per female index patient and led to fewer QALYs lost (0.0272 compared to 0.0308 per index man and 0.0041 compared to 0.0054 per index women). EPT was similarly cost-effective from a societal perspective ($488.34 compared to $592.43 per index man and $193.53 compared to $252.84 per index woman). However, in addition to excluding the QALY impact of complications besides PID, the analysis excluded the potential population transmission effects of the intervention, thus, the accuracy of the findings is uncertain.

### Implications for policy

The results of this model-based economic evaluation indicate that, when compared to standard contact tracing, APT could improve chlamydia-related health outcomes for men and women at a lower total cost. APT should therefore be considered as an alternative to standard contact tracing for chlamydia infection

## Conclusions

Our study found that APT is a cost-effective alternative to standard contact tracing for chlamydia infection in the UK context. The results of this economic evaluation can be used to inform decisions about which PN approach may be the most beneficial in the context of limited healthcare resources. Future work should focus on understanding the timing, duration and probability of complications arising from chlamydia infections, in addition to the health loss from such complications in men.

## Data Availability

The data that support the findings of this study are available from the corresponding author, TR, upon reasonable request.

## Contribution to Authorship

EVW was responsible for the first draft of this manuscript in collaboration with CO. EW carried out the model-based economic analysis. CA carried out the deterministic population-based transmission model. TER oversaw the economic analysis and revised the manuscript. All authors contributed to the editing and revision of the manuscript and gave final approval.

## Details of Ethics Approval

Ethical approval was granted by the UK National Research Ethics Service Committee (London—Chelsea Research Ethics Committee reference: 18/LO/0773).

