## Supplemental Document 1 for "The cost-effectiveness of accelerated partner therapy (APT) compared to standard contact tracing for people with chlamydia: an economic evaluation based on the LUSTRUM population-based chlamydia transmission model"

### Appendix

#### A1 Explanation of costs and resource use

An explanation of how the key costs were calculated and the assumed resource use for each contract tracing strategy and complication is shown in table A1 below:

Table A1 – Details of key costs within the economic models

| Item | Unit Cost | Other information. | Source(s) |
|---|---|---|---|
| *Contract Tracing Costs* | | | |
| APT index patient (notifying partner) | £32.09 | Cost includes a 25-minute consultation (75% band 6 nurse/25% band 7 nurse), 7 minutes of additional staff time (whilst the index patient contacts their sex partner) and one dose of doxycycline (28 x 50mg tablets). | 1,2 |
| APT index patient (not notifying partner) | £22.43 | Cost includes a 25-minute consultation (75% band 6 nurse/25% band 7 nurse) and one dose of doxycycline (28 x 50mg tablets). | 1,2 |
| APT sex partner | £36.17 | Cost includes a 10-minute telephone consultation (75% band 6 nurse/25% band 7 nurse), cost of the telephone call, 10 minutes of administration time, APT pack, chlamydia testing and one dose of doxycycline (28 x 50mg tablets). | 1,2,3 |
| Standard contact tracing index patient | £22.43 | Cost includes a 25-minute consultation (75% band 6 nurse/25% band 7 nurse) and one dose of doxycycline (28 x 50mg tablets). | 1,2 |
| Standard contract tracing sex partner | £38.46 | Cost includes a 30-minute consultation (75% band 6 nurse/25% band 7 nurse), chlamydia testing and one dose of doxycycline (28 x 50mg tablets). | 1,2,3 |
| *Complications* | | | |
| Mild pelvic inflammatory disease | £123.15 | Cost includes a GP visit, chlamydia/HIV testing, one dose of doxycycline (56 x 50mg tablets), one dose of metronidazole (56 x 200mg tablets) and one dose of ceftriaxone (1 x 1g). | 1,2,4,5 |
| Severe pelvic inflammatory disease | £1366.81 | Cost includes 2 GP visits, inpatient stay for 3 days, doxycycline (62 x 50mg tablets), ceftriaxone (3 x 2g) for all women, and an additional laparoscopy for 10% of women | 1,2,4,6 |
| Ectopic pregnancy | £1740.29 | Based on Fourie et al. (2017)[7] it was assumed that 55% of ectopic pregnancies were treated surgically, 30% were treated medically and 15% were treated medically. If managed surgically, cost includes transvaginal ultrasound, salpingectomy, 1 GP visit, urine pregnancy test kit, and an additional clinic visit for 50% of women. If managed medically, cost includes transvaginal ultrasound, methotrexate (medical treatment of ectopic pregnancy), 5 clinic visits, 5 serum hCG tests, 2 liver function tests, full blood count, liver function test and urine pregnancy test kit. If managed expectantly, cost includes transvaginal ultrasound, 5 clinic visits, 5 serum hCG tests, 2 liver function tests, full blood count, liver function test and urine pregnancy test kit. | 1,2,4,5,8 |
| Tubal factor infertility | £3639.06 | Cost includes a GP visit, outpatient visit with a specialist plus follow-up visit, full hormone screen, HIV test, HCV/HCV tests and 1 cycle of IVF (80% fresh cycle and 20% frozen cycle). | 1,4,9,10 |
| Chronic pelvic pain | £652.26 | Cost includes GP visit, outpatient visit with a specialist plus follow-up and transvaginal ultrasound for all women, and an additional laparoscopy for 50% of women. | 1,4 |
| Epididymitis | £187.75 | Cost from Shepherd et al. (2010)[11], inflated to 2019/2020 pounds. | 11 |

**A2 Details of health-state utility value and duration variations for deterministic sensitivity analyses**

The impact of varying both the health-statue utility values (HSUVs) and health state durations from the base-case values was explored using one-way deterministic sensitivity analyses (DSAs). The applied variations are summarised below and are shown in Table A1:

- The HUSVs for the female complications (mild pelvic inflammatory disease (PID), severe PID, ectopic pregnancy, chronic pelvic pain, and tubal factor infertility (TFI)) were varied to the lowest and highest values reported by two previous studies[12,13]
- The HSUV for epididymitis was reduced to 0.45, which is half the value used by the base-case analysis. This arbitrary value HSUV was used in the absence of available studies on health-related quality-of-life for the condition.
- The health state durations for the chronic health states (chronic pelvic pain and TFI) were varied to the lowest and highest values employed by previous cost and cost-effectiveness analyses of STI complications and control interventions.[11,14-16]
- The health state durations for the temporary health states (mild PID, severe PID, ectopic pregnancy and epididymitis) were arbitrarily reduced to half the duration used by the base-case analysis.

**Table A2: Health state utility value and durations variations applied in the deterministic sensitivity analyses.**

| Complication | HSUV | Duration (years) | Source(s) |
|---|---|---|---|
| Mild PID | 0.63-0.91* | 0.0137 | [12,13] |
| Severe PID | 0.57-0.87* | 0.0165 | [12,13] |
| Ectopic pregnancy | 0.58-1.00* | 0.0384 | [12,13] |
| TFI | 0.82-0.87* | 5-15* | [11-13,14] |
| Chronic pelvic pain | 0.60-0.79* | 2-10* | [12,13,16,15] |
| Epididymitis | 0.45 | 0.0151 | - |

Note: * denotes values were drawn from published literature.

**A3 Details and results of deterministic sensitivity analyses**

An extensive range of DSAs were conducted to explore uncertainty in the economic model. The full range of DSAs is summarised below and the results are reported in Table A4a and A4b:

1. The APT sex partner consultation time was reduced from 10 minutes to 6.2 minutes, the average recorded duration from the LUSTRUM RCT.
2. HIV/syphilis tests were included in the APT sex partner packs.
3. The probability of progression from chlamydia infection to PID was reduced to 2% (the lowest probability value in the identified range).
4. The probability of progression from chlamydia infection to PID was increased to 25% (the highest probability value in the identified range).
5. The probability of progression from chlamydia infection to epididymitis was reduced to 1% (the lowest probability value in the identified range).
6. The probability of progression from chlamydia infection to epididymitis was increased to 4% (the highest probability value in the identified range).
7. The probability of progression from PID to ectopic pregnancy was reduced to 2% (the lowest probability value in the identified range).
8. The probability of progression from PID to ectopic pregnancy was increased to 10% (the highest probability value in the identified range).
9. The probability of progression from PID to chronic pelvic pain was reduced to 5% (the lowest probability value in the identified range).
10. The probability of progression from PID to chronic pelvic pain was increased to 20% (the highest probability value in the identified range).
11. The probability of progression from PID to TFI was reduced to 5% (the lowest probability value in the identified range).
12. The probability of progression from PID to TFI was increased to 23% (the highest probability value in the identified range).
13. The probabilities of progression to all complications were simultaneously reduced to the lowest values in the identified range.
14. The probabilities of progression to all complications were simultaneously increased to the highest values in the identified range.
15. The lowest previously published utility values for female complications were used (detailed in Table A2).
16. The highest previously published utility values for female complications were used (detailed in Table A2).

17. The utility value for epididymitis was reduced by 50%
18. The health state durations for mild and severe PID were reduced by 50%
19. The health state duration for epididymitis was reduced by 50%.
20. The health state duration for ectopic pregnancy was reduced by 50%.
21. The health state duration for chronic pelvic pain was reduced to 2 years.
22. The health state duration for chronic pelvic pain was increased to 2 years.
23. The health state duration for TFI was reduced to 5 years.
24. The health state duration for TFI was increased to 10 years.
25. The proportion of women experiencing future pregnancies reduced to 90%. This was 10% lower than the proportion assumed by the base-case analysis as a simplification of the model.
26. Proportion of women with TFI undergoing IVF reduced to 50%. This was 50% lower than the proportion assumed by the base-case analysis as a simplification of the model.
27. All ectopic pregnancies were assumed to be managed surgically.

**Table A3.1 – Deterministic sensitivity analysis results**

| **1. APT sex partner consultations reduced to 6.2 minutes** | | | | | | |
|---|---|---|---|---|---|---|
| Contract Tracing Approach | Intervention Cost (£) | Total Cost (£) | MOs | QALYs lost | ICER/MOA (£) | ICER/ QALY gained (£) |
| APT | 128,453 | 363,909 | 518 | 181 | Dominant | Dominant |
| Standard Contract Tracing | 116,334 | 379,597 | 591 | 202 | - | - |
| **2. HIV/syphilis tests included in APT sex partner packs/as tests for standard contract tracing.** | | | | | | |
| Contract Tracing Approach | Intervention Cost (£) | Total Cost (£) | MOs | QALYs lost | ICER/MOA (£) | ICER/ QALY gained (£) |
| APT | 147,510 | 382,967 | 518 | 181 | Dominant | Dominant |
| Standard Contract Tracing | 125,864 | 389,127 | 591 | 202 | - | - |
| **3. PID progression reduced to 2%** | | | | | | |
| Contract Tracing Approach | Intervention Cost (£) | Total Cost (£) | MOs | QALYs lost | ICER/MOA (£) | ICER/ QALY gained (£) |
| APT | 135,201 | 191,040 | 149 | 37 | 593 | 2797 |
| Standard Contract Tracing | 116,334 | 178,862 | 169 | 41 | - | - |
| **4. PID progression increased to 25%** | | | | | | |
| Contract Tracing Approach | Intervention Cost (£) | Total Cost (£) | MOs | QALYs lost | ICER/MOA (£) | ICER/ QALY gained (£) |
| APT | 135,201 | 713,976 | 1224 | 456 | Dominant | Dominant |
| Standard Contract Tracing | 116,334 | 763,280 | 1398 | 510 | - | - |

**Table A3.2 – Deterministic sensitivity analysis results (continued)**

| | | | | | | |
|---|---|---|---|---|---|---|
| **5. Epididymitis progression reduced to 1%** | | | | | | |
| Contract Tracing Approach | Intervention Cost (£) | Total Cost (£) | MOs | QALYs lost | ICER/MOA (£) | ICER/ QALY gained (£) |
| APT | 135,201 | 365,474 | 490 | 181 | Dominant | Dominant |
| Standard Contract Tracing | 116,334 | 373,742 | 560 | 202 | - | - |
| **6. Epididymitis progression increased to 4%** | | | | | | |
| Contract Tracing Approach | Intervention Cost (£) | Total Cost (£) | MOs | QALYs lost | ICER/MOA (£) | ICER/QALY gained (£) |
| APT | 135,201 | 381,023 | 573 | 181 | Dominant | Dominant |
| Standard Contract Tracing | 116,334 | 391,306 | 654 | 202 | - | - |
| **7. Ectopic pregnancy progression reduced to 2%** | | | | | | |
| Contract Tracing Approach | Intervention Cost (£) | Total Cost (£) | MOs | QALYs lost | ICER/MOA (£) | ICER/ QALY gained (£) |
| APT | 135,201 | 361,370 | 513 | 181 | Dominant | Dominant |
| Standard Contract Tracing | 116,334 | 369,218 | 585 | 202 | - | - |
| **8. Ectopic pregnancy progression increased to 10%** | | | | | | |
| Contract Tracing Approach | Intervention Cost (£) | Total Cost (£) | MOs | QALYs lost | ICER/MOA (£) | ICER/QALY gained (£) |
| APT | 135,201 | 398,517 | 534 | 181 | Dominant | Dominant |
| Standard Contract Tracing | 116,334 | 410,732 | 609 | 203 | - | - |
| **9. Chronic pelvic pain progression reduced to 5%** | | | | | | |
| Contract Tracing Approach | Intervention Cost (£) | Total Cost (£) | MOs | QALYs lost | ICER/MOA (£) | ICER/ QALY gained (£) |
| APT | 135,201 | 355,682 | 495 | 131 | Dominant | Dominant |
| Standard Contract Tracing | 116,334 | 362,861 | 559 | 146 | - | - |
| **10. Chronic pelvic pain progression increased to 20%** | | | | | | |
| Contract Tracing Approach | Intervention Cost (£) | Total Cost (£) | MOs | QALYs lost | ICER/MOA (£) | ICER/QALY gained (£) |
| APT | 135,201 | 387,771 | 544 | 238 | Dominant | Dominant |
| Standard Contract Tracing | 116,334 | 398,723 | 629 | 266 | - | - |
| **11. TFI progression reduced to 5%** | | | | | | |
| Contract Tracing Approach | Intervention Cost (£) | Total Cost (£) | MOs | QALYs lost | ICER/MOA (£) | ICER/ QALY gained (£) |
| APT | 135,201 | 333,133 | 508 | 140 | Dominant | Dominant |
| Standard Contract Tracing | 116,334 | 337,661 | 580 | 157 | - | - |

**Table A3.3 – Deterministic sensitivity analysis results (continued)**

| | | | | | | |
|---|---|---|---|---|---|---|
| **12. TFI progression increased to 23%** | | | | | | |
| Contract Tracing Approach | Intervention Cost (£) | Total Cost (£) | MOs | QALYs lost | ICER/MOA (£) | ICER/QALY gained (£) |
| APT | 135,201 | 501,992 | 554 | 324 | Dominant | Dominant |
| Standard Contract Tracing | 116,334 | 526,373 | 632 | 362 | - | - |
| **13. All complications reduced to lowest probability in range (PID = 2%, TFI = 5%, ectopic pregnancy = 2%, chronic pelvic pain = 5%, epididymitis = 1%)** | | | | | | |
| Contract Tracing Approach | Intervention Cost (£) | Total Cost (£) | MOs | QALYs lost | ICER/MOA (£) | ICER/ QALY gained (£) |
| APT | 135,201 | 173,375 | 113 | 18 | 977 | 6639 |
| Standard Contract Tracing | 116,334 | 159,058 | 128 | 20 | - | - |
| **14. All complications increased to highest probability in range (PID = 25%, TFI = 23%, ectopic pregnancy = 10%, chronic pelvic pain = 20%, epididymitis = 4%)** | | | | | | |
| Contract Tracing Approach | Intervention Cost (£) | Total Cost (£) | MOs | QALYs lost | ICER/MOA (£) | ICER/QALY gained (£) |
| APT | 135,201 | 1,169,566 | 1477 | 963 | Dominant | Dominant |
| Standard Contract Tracing | 116,334 | 1,272,557 | 1702 | 1077 | - | - |
| **15. Proportion of severe PID increased to 25%** | | | | | | |
| Contract Tracing Approach | Intervention Cost (£) | Total Cost (£) | MOs | QALYs lost | ICER/MOA (£) | ICER/QALY gained (£) |
| APT | 135,201 | 443,324 | 518 | 181 | Dominant | Dominant |
| Standard Contract Tracing | 116,334 | 460,808 | 591 | 202 | - | - |
| **16. HSUVs from the literature (lower bound)** | | | | | | |
| Contract Tracing Approach | Intervention Cost (£) | Total Cost (£) | MOs | QALYs lost | ICER/MOA (£) | ICER/ QALY gained (£) |
| APT | 135,201 | 370,657 | 518 | 103 | Dominant | Dominant |
| Standard Contract Tracing | 116,334 | 379,597 | 591 | 115 | - | - |
| **17. HSUVs from the literature (upper bound)** | | | | | | |
| Contract Tracing Approach | Intervention Cost (£) | Total Cost (£) | MOs | QALYs lost | ICER/MOA (£) | ICER/ QALY gained (£) |
| APT | 135,201 | 370,657 | 518 | 60 | Dominant | Dominant |
| Standard Contract Tracing | 116,334 | 379,597 | 591 | 67 | - | - |
| **18. Utility value for epididymitis reduced by 50%** | | | | | | |
| Contract Tracing Approach | Intervention Cost (£) | Total Cost (£) | MOs | QALYs lost | ICER/MOA (£) | ICER/ QALY gained (£) |
| APT | 135,201 | 370,657 | 518 | 181 | Dominant | Dominant |
| Standard Contract Tracing | 116,334 | 379,597 | 591 | 203 | - | - |

**Table A3.4 – Deterministic sensitivity analysis results (continued)**

| Contract Tracing Approach | Intervention Cost (£) | Total Cost (£) | MOs | QALYs lost | ICER/MOA (£) | ICER/ QALY gained (£) |
|---|---|---|---|---|---|---|
| **19. PID health state duration halved** | | | | | | |
| Contract Tracing Approach | Intervention Cost (£) | Total Cost (£) | MOs | QALYs lost | ICER/MOA (£) | ICER/ QALY gained (£) |
| APT | 135,201 | 370,657 | 518 | 179 | Dominant | Dominant |
| Standard Contract Tracing | 116,334 | 379,597 | 591 | 201 | - | - |
| **20. Epididymitis health state duration halved** | | | | | | |
| Contract Tracing Approach | Intervention Cost (£) | Total Cost (£) | MOs | QALYs lost | ICER/MOA (£) | ICER/QALY gained (£) |
| APT | 135,201 | 370,657 | 518 | 181 | Dominant | Dominant |
| Standard Contract Tracing | 116,334 | 379,597 | 591 | 202 | - | - |
| **21. Ectopic pregnancy health state duration halved** | | | | | | |
| Contract Tracing Approach | Intervention Cost (£) | Total Cost (£) | MOs | QALYs lost | ICER/MOA (£) | ICER/ QALY gained (£) |
| APT | 135,201 | 370,657 | 518 | 181 | Dominant | Dominant |
| Standard Contract Tracing | 116,334 | 379,597 | 591 | 202 | - | - |
| **22. CPP health state duration reduced to 2 years** | | | | | | |
| Contract Tracing Approach | Intervention Cost (£) | Total Cost (£) | MOs | QALYs lost | ICER/MOA (£) | ICER/QALY gained (£) |
| APT | 135,201 | 370,657 | 518 | 159 | Dominant | Dominant |
| Standard Contract Tracing | 116,334 | 379,597 | 591 | 176 | - | - |
| **23. CPP health state duration increased to 10 years** | | | | | | |
| Contract Tracing Approach | Intervention Cost (£) | Total Cost (£) | MOs | QALYs lost | ICER/MOA (£) | ICER/ QALY gained (£) |
| APT | 135,201 | 370,657 | 518 | 281 | Dominant | Dominant |
| Standard Contract Tracing | 116,334 | 379,597 | 591 | 315 | - | - |
| **24. TFI health state duration reduced to 5 years** | | | | | | |
| Contract Tracing Approach | Intervention Cost (£) | Total Cost (£) | MOs | QALYs lost | ICER/MOA (£) | ICER/QALY gained (£) |
| APT | 135,201 | 370,657 | 518 | 144 | Dominant | Dominant |
| Standard Contract Tracing | 116,334 | 379,597 | 591 | 161 | - | - |
| **25. TFI health state duration increased to 15 years** | | | | | | |
| Contract Tracing Approach | Intervention Cost (£) | Total Cost (£) | MOs | QALYs lost | ICER/MOA (£) | ICER/ QALY gained (£) |
| APT | 135,201 | 370,657 | 518 | 216 | Dominant | Dominant |
| Standard Contract Tracing | 116,334 | 379,597 | 591 | 242 | - | - |
| **26. 90% of women considering future pregnancy** | | | | | | |
| Contract Tracing Approach | Intervention Cost (£) | Total Cost (£) | MOs | QALYs lost | ICER/MOA (£) | ICER/QALY gained (£) |
| APT | 135,201 | 360,357 | 515 | 171 | Dominant | Dominant |
| Standard Contract Tracing | 116,334 | 368,086 | 588 | 192 | - | - |

**Table A3.5 – Deterministic sensitivity analysis results (continued)**

| **27. 50% of women TFI undergoing IVF** | | | | | | |
|---|---|---|---|---|---|---|
| Contract Tracing Approach | Intervention Cost (£) | Total Cost (£) | MOs | QALYs lost | ICER/MOA (£) | ICER/ QALY gained (£) |
| APT | 135,201 | 333,134 | 518 | 181 | Dominant | Dominant |
| Standard Contract Tracing | 116,334 | 337,668 | 591 | 202 | - | - |
| **28. All ectopic pregnancies treated surgically** | | | | | | |
| Contract Tracing Approach | Intervention Cost (£) | Total Cost (£) | MOs | QALYs lost | ICER/MOA (£) | ICER/QALY gained (£) |
| APT | 135,201 | 376,053 | 518 | 181 | Dominant | Dominant |
| Standard Contract Tracing | 116,334 | 385,627 | 591 | 202 | - | - |

**A4 Robustness checks**

Robustness checks were made to ensure correct functioning of the model. For example, the probability of progression from chlamydia infection to PID was set to 0% to determine if the economic model output followed intuition, i.e., APT would not be cost-effective. The results, presented in Table A5, indicate correct functioning of the model as the incremental cost-effectiveness ratio in terms of quality-adjusted life years gained (£814,148/QALY) significantly exceeds the NICE recommended threshold of £20,000 - £30,000 per QALY gained.

**Table A4 – Robustness check using 0% probability of progression from chlamydia infection to PID.**

| **Contract Tracing Approach** | **Intervention Cost (£)** | **Total Cost (£)** | **MOs** | **QALYs lost** | **ICER/MOA (£)** | **ICER/QALY (£)** |
|---|---|---|---|---|---|---|
| APT | 135,201 | 145,567 | 55 | 0.166 | 2451 | 814,148 |
| Standard Contract Tracing | 116,334 | 128,043 | 62 | 0.188 | - | - |
